# Frailty variation models for susceptibility and exposure to SARS-CoV-2

**DOI:** 10.1101/2021.05.25.21257766

**Authors:** M. Gabriela M. Gomes, Marcelo U. Ferreira, Maria Chikina, Wesley Pegden, Ricardo Aguas

## Abstract

Individual variation in susceptibility and exposure is subject to selection by force of infection, accelerating the natural acquisition of immunity, and reducing herd immunity thresholds and epidemic final sizes. This is a manifestation of a wider population phenomenon known as “frailty variation” in demography. Despite this theoretical understanding, public health policies continue to be guided by mathematical models that leave out most of the relevant variation and as a result inflate projected infection burdens. Here we focus on the trajectories of the coronavirus disease (COVID-19) pandemic in England and Scotland. We fit models to series of daily deaths and estimate relevant epidemiological parameters, including coefficients of variation which we find in agreement with direct measurements based on published contact surveys. Our estimates are robust to whether the data series encompass one or two pandemic waves. We conclude that herd immunity thresholds are being reached with a larger contribution of vaccination in Scotland than in England, where naturally acquired immunity is higher. These results are relevant to global vaccination policies.

## 1. Introduction

Almost 100 years ago, Kermack and McKendrick (Kermack and McKendrick 1927) and McKendrick (McKendrick 1939) fitted susceptible-infected-recovered (SIR) models to observed epidemics and alerted for the simplifying assumption “that all infected persons are equivalent, and that all susceptible persons are equally liable to acquire infection” (McKendrick 1939). In their fittings they adjust not only transmission parameters but also the size of the susceptible and exposed population at epidemic onset. Susceptible and exposed population sizes needed to be adjusted so their homogeneous models could fit the data.

Thirty years later, Gart (Gart 1968) admited that “it is difficult to define exactly the size of the population of susceptible hosts. In this instance the difficulty is associated with the heterogeneous nature of the population”. The author divided the population in two groups, depending on their history of infection, and allowed much greater susceptibility in the group with no history. This did not seem sufficient to provide good fit to observed epidemics as the author adds “we assume that the first group is a homogeneous group of susceptibles, while the second is actually a mixture of immune and susceptible individuals”. In (Gart 1971) the author extend the model to several susceptibility groups, and, more than a decade later decade later, (Ball 1985) compared a model with several susceptibility groups with the homogeneous version and described how homogeneity increases epidemic size. (Coutinho *et al*. 1999) expand the formalisms but conclude that “at present practical applications might be difficult”. (Pastor-Satorras and Vespignani 2001) developed related formalisms to describe epidemics on contact networks.

Meanwhile, frailty variation had been formalised in demography (Vaupel *et al*. 1979) and introduced in practice in survival analysis (Aalen 1988) and non-communicable disease epidemiology (Aalen *et al*. 2015) to improve model fits and interpretation.

On the experimental front, (Dwyer *et al*. 1997) measured nonlinear relationships between transmission and densities of susceptible hosts, implying that the bilinear term in the classical SIR model may not be appropriate. The authors attributed this non-linearity in transmission to heterogeneity in host susceptibility to infection which they estimated from the shapes of dose-response curves.

(Finkenstadt and Grenfell 2000) fitted a model with nonlinear relationships between transmission and density of susceptible hosts to an observed epidemic and estimated the exponent which they interpreting as representing heterogeneity in mixing. (Novozhilov 2008) derived the expressions for the exponents from explicit distributions of susceptibility.

Here we build on this history and analyse the coronavirus disease (COVID-19) with frailty variation models. The study is focused on England and Scotland, where infection was first detected in early 2020.

We use susceptible-exposed-infected-recovered (SEIR) models (Diekmann *et al*. 2013) incorporating distributions of individual susceptibility or exposure to infection (Ball 1985; Coutinho *et al*. 1999; Gomes *et al*. 2020; Katriel 2012; Novozhilov 2008). We use Bayesian inference to estimate the model parameters by fitting series of deaths while accounting for the combined effects of non-pharmaceutical interventions (NPIs), behavioural change, seasonality and viral evolution. We estimate coefficients of variation which are in agreement with direct measurements based on contact-pattern studies, e.g., (Mossong *et al*. 2008; Hens *et al*. 2009; Willem *et al*. 2012). We show that individual variation in susceptibility or exposure to infection can significantly affect model projections and should be accounted for to describe the dynamics of SARS-CoV-2.

## 2. Mathematical models

The basic compartmental SEIR model describing the transmission dynamics of SARS-CoV-2 is represented diagrammatically in Fig. 1. Following (Gomes *et al*. 2020) the model accounts for individual variation in susceptibility or exposure to infection.

**Fig. 1.**
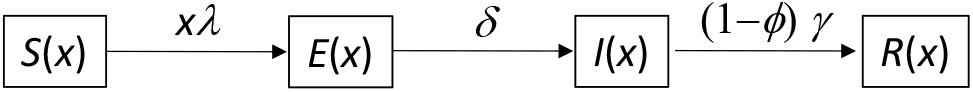
Susceptible-exposed-infected-recovered (SEIR) compartmental model representing the transmission dynamics of SARS-CoV-2 in a heterogeneous host population.

### 2.1. Individual variation in susceptibility to infection

Let *x* denote the individual susceptibility to infection in relation to the mean, which we describe by a gamma distribution *q*(*x*) with mean ∫ *xq*(*x*)*dx* = 1 and parametrised by a coefficient of variation, 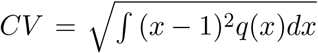. Susceptible individuals, *S*(*x*), become exposed at a rate that depends on their susceptibility *x* and on the average force of infection *λ* which accounts for the total number of infectious individuals in the population over time. Upon exposure, susceptible individuals enter an incubation phase, *E*(*x*), during which they gradually become infectious (Wei *et al*. 2020; To *et al*. 2020; Arons *et al*. 2020; He *et al*. 2020). The infectiousness in this phase is made to be half that in the following stage (*ρ* = 0.5), to which individuals transit within an average of 5.5 days (*δ* = 1*/*5.5) (McAloon *et al*. 2020). The fully infectious state is denoted by *I*(*x*). Infected individuals are eventually removed, on average approximately 4 days after becoming fully infectious (*γ* = 1*/*4) (Nishiura *et al*. 2020; Lauer *et al*. 2020; Li *et al*. 2020). A small fraction ***ϕ***(*x*) die to COVID-19 while the remaining majority recover into *R*(*x*) where they are noninfectious and temporarily resistant to reinfection due to acquired immunity. The model is represented diagrammatically in Fig. 1 and mathematically by the infinite system of ordinary differential equations:

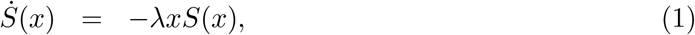

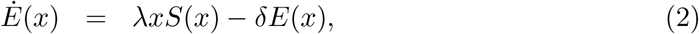

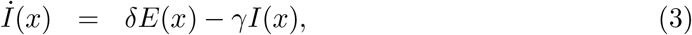

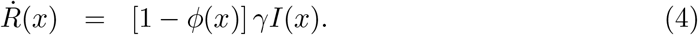

The average force of infection upon susceptible individuals in a population of size *N* and transmission coefficient *β* is defined by:

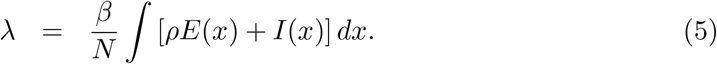

An epidemic is simulated by introducing a small seed of infectious individuals in a susceptible population. Initial growth of infected numbers is near exponential but decelerates as individuals are removed from the susceptible pool by infection and immunity. With variation in susceptibility, highly susceptible individuals tend to be infected earlier, leaving behind a residual pool of lower mean susceptibility. This selective depletion intensifies the deceleration of epidemic growth and gives an efficient head start to the acquisition of population immunity. Eventually the epidemic will subside and the herd immunity threshold, defining the percentage of the population that needs to be immune to reverse epidemic growth and prevent future waves, is lower when variation in susceptibility is higher.

The basic reproduction number, defined as the number of infections caused by an average infected individual in a totally susceptible population, is written for system (1)-(4) with force of infection (5) as:

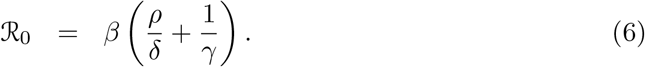

This is a crude indicator of early transmissibility but its use quickly becomes cumbersome. Several factors, such as NPIs, human behaviour, seasonality and viral evolution, affect ℛ_0_ in a time-dependent manner. We denote the resulting quantity by ℛ_c_(*t*) = *c*(*t*) ℛ_0_, where *c*(*t*) *>* 0 describes the basic risk of infection at time *t* in relation to baseline.

In the estimation of ℛ_c_(*t*) we assume a profile for *c*(*t*) as illustrated in Fig. 2: *T*_0_ is the time when ℛ_0_ begins to show decrease due to behavioural change or seasonality; *L*_1_ is the period of maximal contact restrictions due to lockdown (48 days in England, from 26 March to 12 May, and 66 days in Scotland, from 24 March to 28 May, 2020) and *c*_1_ ≤ 1 is the value of ℛ_c_(*t*) during *L*_1_ in relation to the initial ℛ_0_; *T*_1_ is the time elapsed between *T*_0_ and *L*_1_, over which transmission is allowed to decrease linearly. After *L*_1_, contact restrictions are progressively relaxed and we allow transmission to begin a linear increase such that *c*(*t*) reaches 1 in *T*_2_ days, which may or may not be within the range of the study. Changes in other factors that affect transmission (such as seasonality or viral evolution) are inseparable from contact changes in this framework and are also accounted for by *c*(*t*).

**Fig. 2.**
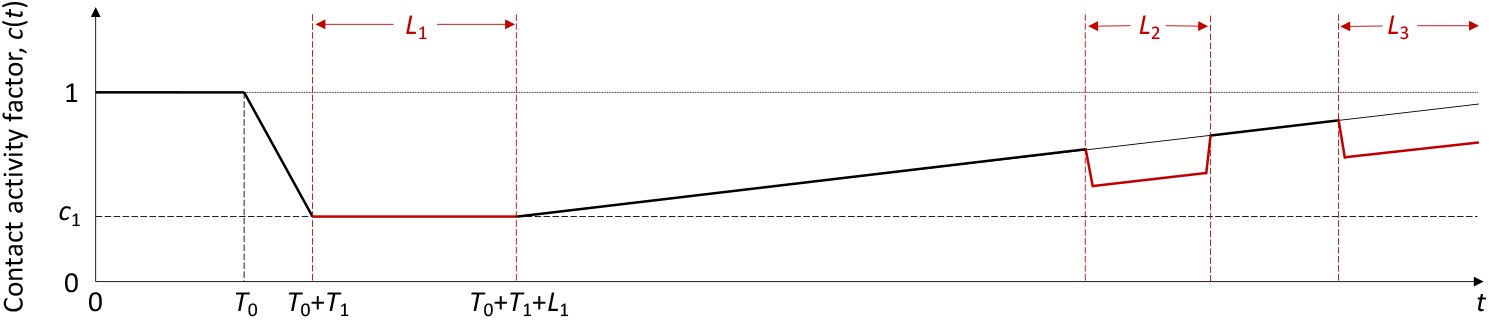
Schematic illustration of factor *c*(*t*) representing the combined effects of NPIs, seasonality and viral evolution on the reproduction number.

The model will be used to analyse COVID-19 deaths recorded over approximately one year. Mathematically this is constructed as:

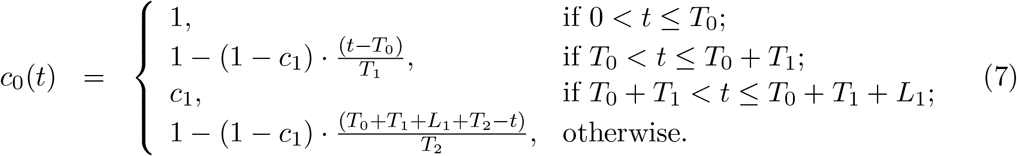

Second and third lockdowns in the autumn and winter season are implemented more simply as a further reduction in transmission (by a factor *c*_2_) over the stipulated time periods. Specifically:

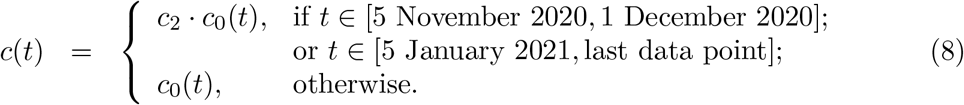

A finite version of system (1)-(4) and (5) can be derived exactly (Novozhilov 2008):

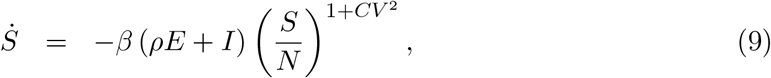

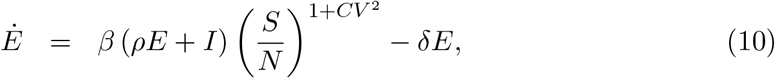

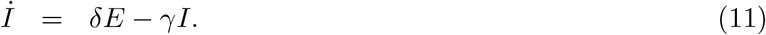

If ℛ_0_ had remained constant throughout the duration of the study, a herd immunity threshold (HIT) would be derived as in (Montalb’san *et al*. 2020):

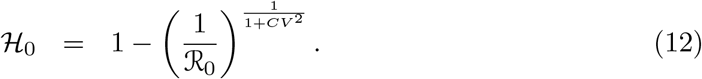

The expectation, however, is that ℛ_0_ changes due to seasonal effects and viral evolution. These effects are currently inseparable from those of NPIs and behavioural change and, consequently, we cannot obtain a time-depend ℛ_0_. Although our results for variable susceptibility models will be accompanied by HIT estimates calculated according to formula (12) we highlight that these refer to a virus as transmissible as the SARS-CoV-2 of early 2020. Once reliable estimates are available for evolving transmissibility, HIT estimates can be updated.

In reality, as infection spreads, the susceptible compartment *S* is depleted and recovered individuals populate compartment *R* where they are protected by acquired immunity. Eventually they lose that protection as immunity wanes or is evaded by new viral variants. This is omitted in this version of the model given our purpose to analyse data reported over one year when the frequency of reinfection has been relatively low in our study setting (Hall *et al*. 2021). In Supplementary Information, however, we formulate an extended model with reinfection to show that the addition does not change our conclusions.

### 2.2. Individual variation in exposure to infection

In a directly transmitted infectious disease, such as COVID-19, variation in exposure to infection is primarily governed by patterns of connectivity among individuals. We incorporate this in system (1)-(4) assuming that individuals mix at random (Pastor-Satorras and Vespignani 2001; Miller *et al*. 2012). In a separate study we developed an assortative mixing version of the model adopted here and did not find the results of interest to change (Aguas *et al*. 2020). Under random mixing and heterogeneous connectivity, the force of infection is written as

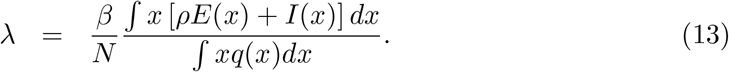

The basic reproduction number is

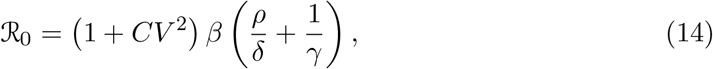

and ℛ_c_(*t*) = *c*(*t*) · ℛ_0_ is as above.

As with variable susceptibility, model (1)-(4) with variable exposure (13) can be reduced to a 3-dimensional system of ODEs:

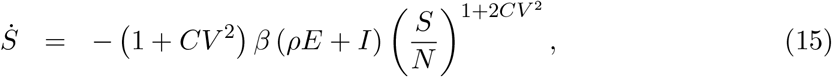

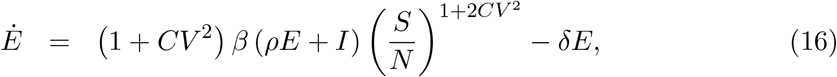

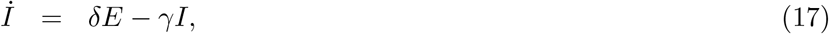

the effective reproduction number written as:

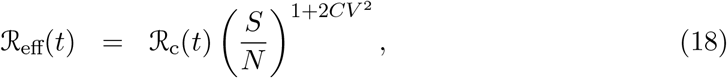

and the herd immunity threshold derived as in (Montalb’san *et al*. 2020):

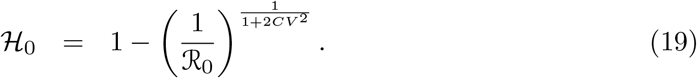

## 3. Data

We used publicly available epidemiological data from the UK coronavirus dashboard [https://coronavirus.data.gov.uk/] describing the unfolding of the SARS-CoV-2 epidemic, to estimate relevant transmission related parameters for the larger nations: England (56 million inhabitants) and Scotland (5.5 million). Namely, we collected datasets containing daily deaths (deaths within 28 days of positive test by date of death), 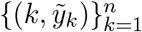, where *k* = 1 is the day when the cumulative moving average of death numbers exceeded 5 · 10^−8^ of the population in both nations (10 March 2020).

Model outputs would then be fitted to the raw series of daily deaths between 10 March 2020 and 1 February 2021 (*n* = 329 days in total) adopting an infection fatality ratio of 0.9% (Ward *et al*. 2021) throughout the study period and initial conditions:

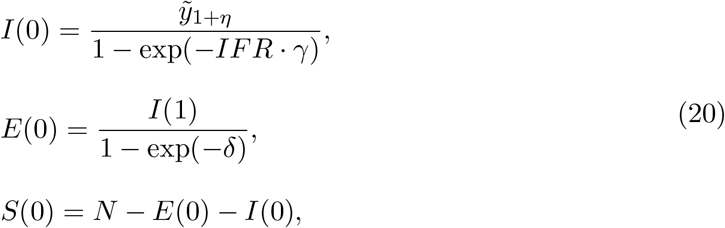

where *η* is the excess duration of a fatal infection relatively to non-fatal. In Supplementary Information we explore the sensitivity of the heterogeneous susceptibility results to changing the value of IFR.

## 4. Model fitting and parameter estimation

In order to preserve identifiability, we made five simplifying assumptions: (i) the infection fatality ratio (IFR) is constant throughout the study period; (ii) natural (seasonality and viral evolution) and interventional (NPI) modulators of the reproduction number are encapsulated into a single time varying parameter *c*(*t*) as illustrated in Fig. 2; (iii) excess transmission from critically ill stages is negligible; (iv) reinfection is negligible; (v) vaccine effects are negligible during the fitted period, which in section 5.1 ends on 1 February 2021 (less than 1% fully vaccinated in the UK) and in section 5.2 ends on 1 July 2020 (no vaccines were in use).

Parameter estimation was performed with the software MATLAB (MathWorks, Natick, MA) using the PESTO (Parameter EStimation TOolbox) package (Stapor *et al*. 2018). We assumed that the number of SARS-CoV-2 infections are Poisson distributed.

We try to reproduce the dynamics of COVID-19 deaths by estimating the set of parameters *θ* that maximises the log-likelihood (LL) of observing the daily numbers of reported deaths *Y* :

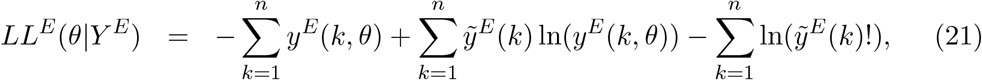

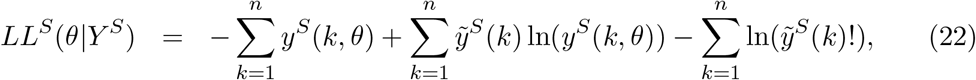

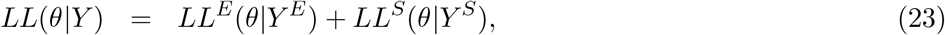

in which *y*^*E*^(*k, θ*) and *y*^*S*^(*k, θ*) are the simulated model output numbers of COVID-19 deaths at day *k* in England and Scotland for the set of parameters *θ*, 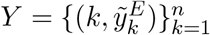 and 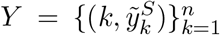 are the numbers of daily reported deaths, and *n* is the total number of days included in the analysis.

The set of parameters to be estimated is:

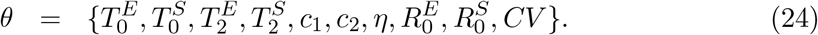

To ensure that the estimated maximum is a global maximum, we performed 50 multistart optimisations with initialisation parameters sampled from a Latin-Hypercube. The combination of parameters resulting in the maximal log-likelihood were used as a starting point for 100, 000 Markov Chain Monte-Carlo iterations. From the resulting posterior distributions, we extracted the median estimates for each parameter and the respective 95% credible intervals. We used uniformly distributed priors with wide ranges.

We apply the outlined fitting procedure using both heterogeneity models (specifically individual variation in susceptibility to infection and individual variation in exposure to infection) as well as a homogeneity model where we set the coefficient of variation to zero (*CV* = 0). The Akaike information criterion (AIC) is then applied to select the best fitting model.

## 5. Results and Discussion

### 5.1. Estimated parameters and herd immunity thresholds

Variable susceptibility, variable connectivity and homogeneous models were fitted to series of COVID-19 death reported in England and Scotland until 1 February 2021. The fits are shown in Figs. 3, 4 and 5 (fitted data points in green), and the estimated parameters in Table 1. Maximum log-likelihoods are also displayed, as well as AIC scores for model selection. We conclude that variable susceptibility and variable connectivity models are better supported by the data (lower AIC in bold) than the homogeneous model\as found previously (Aguas *et al*. 2020; Colombo *et al*. 2020). Variable connectivity is slightly better supported although the reality most likely combines the two forms of heterogeneity.

**Table 1.** Model parameters estimated by Bayesian inference based on daily deaths until 1 February 2021. Model selection based on maximum log-likelihood (LL) and Akaike information criterion (AIC). Best fitting models have lower AIC scores. Infection fatality ratio, IFR = 0.9%. ℋ_0_, calculated from ℛ_0_ and *CV* using formulas (12) or (19), as appropriate.

|  | Heterogeneous susceptibility |  | Heterogeneous connectivity |  | Homogeneous |  |
| --- | --- | --- | --- | --- | --- | --- |
|  | Median | 95% CI | Median | 95% CI | Median | 95% CI |
| *Common parameters* |  |  |  |  |  |  |
| $c_1$ | 0.25 | (0.24, 0.26) | 0.24 | (0.24, 0.25) | 0.21 | (0.21, 0.21) |
| $c_2$ | 0.68 | (0.67, 0.73) | 0.65 | (0.64, 0.67) | 0.57 | (0.57, 0.57) |
| $\eta$ | 11 | (11, 11) | 12 | (12, 12) | 8 | (8, 8) |
| $CV$ | 1.59 | (1.58, 1.62) | 1.14 | (1.12, 1.17) | 0 | — |
| *England* |  |  |  |  |  |  |
| $T_0$ | 5.13 | (5.08, 5.61) | 3.92 | (3.87, 3.95) | 4.56 | (4.56, 4.58) |
| $T_2$ | 398.90 | (396.50, 400.14) | 389.50 | (382.44, 393.46) | 685.98 | (685.47, 686.80) |
| $\mathcal{R}_0$ | 3.41 | (3.35, 3.45) | 3.47 | (3.46, 3.48) | 3.67 | (3.67, 3.68) |
| $\mathcal{H}_0$ | 29% | (28%, 30%) | 29% | (28%, 30%) | 73% | (73%, 73%) |
| *Scotland* |  |  |  |  |  |  |
| $T_0$ | 8.53 | (8.27, 9.53) | 8.96 | (8.43, 9.61) | 9.07 | (9.03, 9.09) |
| $T_2$ | 570.38 | (557.91, 581.11) | 518.79 | (500.88, 532.28) | 847.43 | (845.30, 848.62) |
| $\mathcal{R}_0$ | 3.72 | (3.60, 3.77) | 3.72 | (3.68, 3.74) | 3.96 | (3.96, 3.97) |
| $\mathcal{H}_0$ | 31% | (30%, 32%) | 31% | (30%, 31%) | 75% | (75%, 75%) |
| *Model selection* |  |  |  |  |  |  |
| LL | −4214 |  | −3712 |  | −6079 |  |
| AIC | 8448 |  | **7443** |  | 12176 |  |

**Fig. 3.**
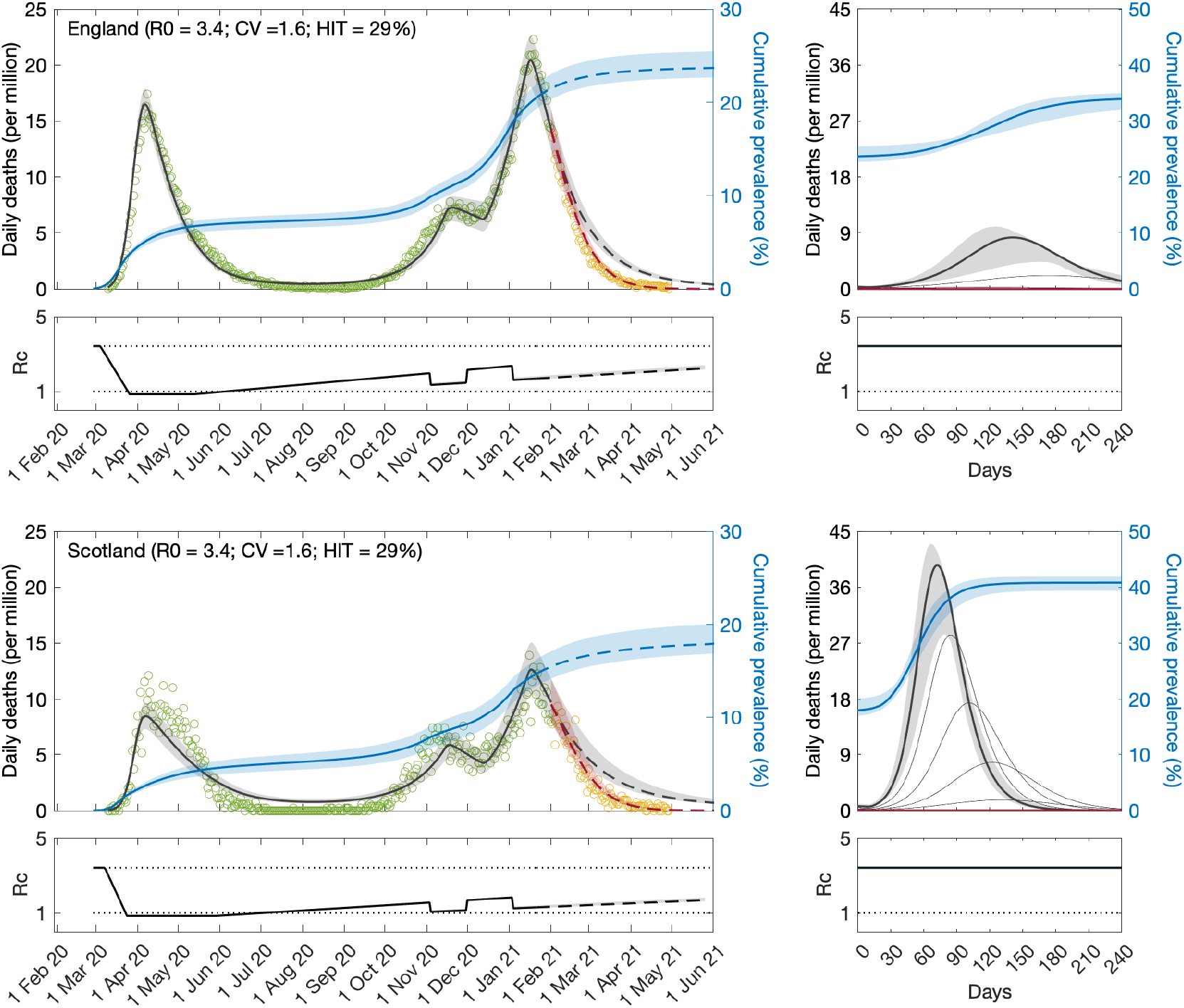
SARS-CoV-2 transmission in England and Scotland with individual variation in susceptibility to infection. Susceptibility factors implemented as gamma distributions. Modelled trajectories of COVID-19 deaths (black and red curves) and cumulative percentage infected (blue). Dots are data for daily reported deaths: fitted (green); posterior to fitted time period (yellow). Basic reproduction numbers under control (ℛ_c_) are displayed on shallow panels underneath the main plots. Left panels represent fitted segments as solid curves and projected scenarios as dashed: without vaccination (black); with a vaccination programme that effectively immunises 8% of the unvaccinated population per month from February onwards (red). Right panels prolong those projections further in time assuming ℛ_c_(*t*) = ℛ_0_ (heavier curves) and explore additional vaccination scenarios (thin curves), from top to bottom (in % of unvaccinated population per month): 0.5, 1, 1.5, 2, 3, 4, 5, 6, 7 (black); 9, 10, 11, 12, 13, 14, 15, 16 (red). Inputed parameter values: *δ* = 1*/*4 per day; *γ* = 1*/*5.5 per day; *ρ* = 0.5; and infection fatality ratio IFR = 0.9%. Inicial basic reproduction numbers, coefficients of variation and control parameters estimated by Bayesian inference (estimates in Table 1). Fitted curves represent best fitting trajectories and shades are 95% credible intervals generated from 100, 000 posterior samples.

**Fig. 4.**
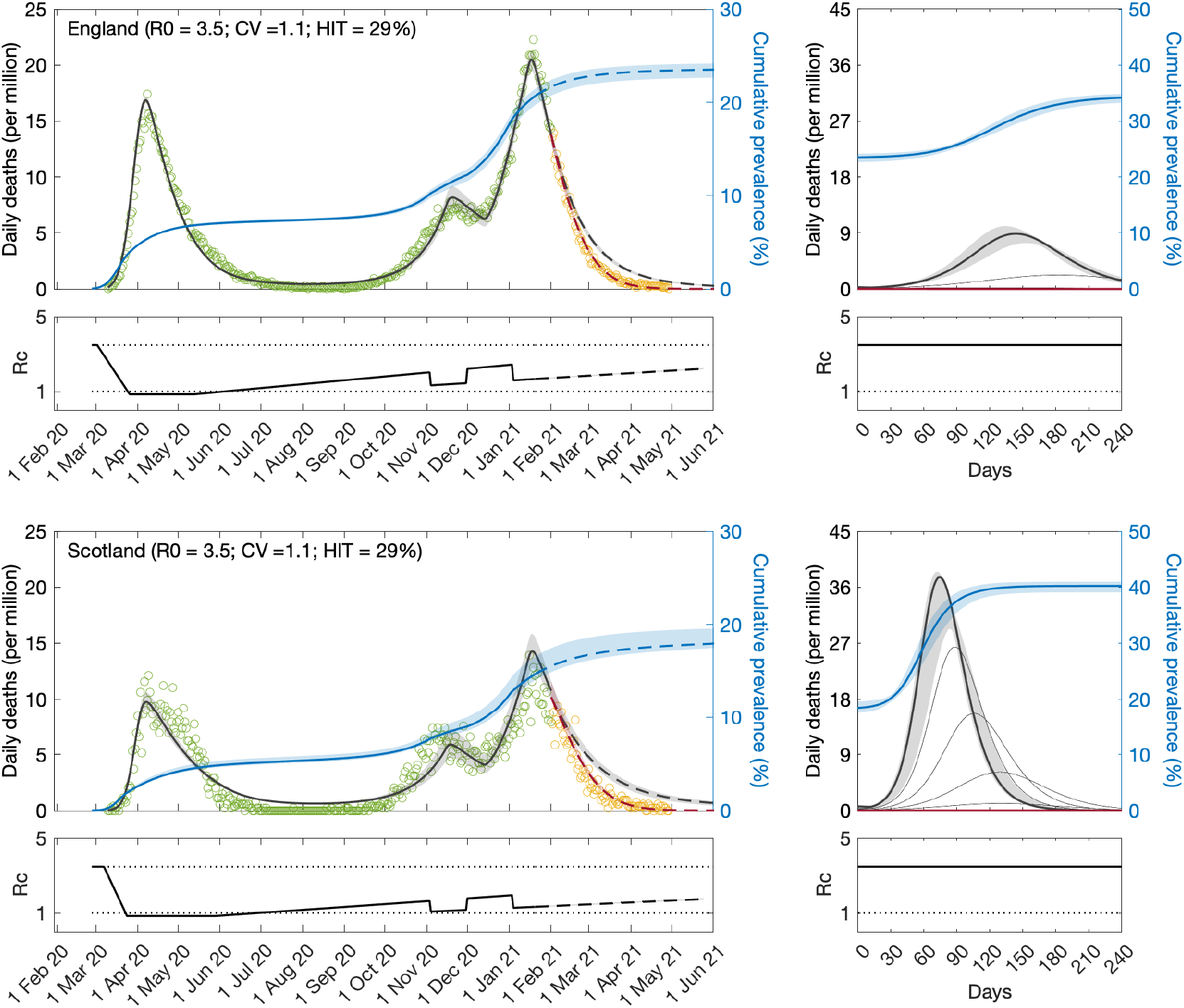
SARS-CoV-2 transmission in England and Scotland with individual variation in exposure to infection. Connectivity factors implemented as gamma distributions. Modelled trajectories of COVID-19 deaths (black and red curves) and cumulative percentage infected (blue). Dots are data for daily reported deaths: fitted (green); posterior to fitted time period (yellow). Basic reproduction numbers under control (ℛ_c_) are displayed on shallow panels underneath the main plots. Left panels represent fitted segments as solid curves and projected scenarios as dashed: without vaccination (black); with a vaccination programme that effectively immunises 8% of the unvaccinated population per month from February onwards (red). Right panels prolong those projections further in time assuming ℛ_c_(*t*) = ℛ_0_ (heavier curves) and explore additional vaccination scenarios (thin curves), from top to bottom (in % of unvaccinated population per month): 0.5, 1, 1.5, 2, 3, 4, 5, 6, 7 (black); 9, 10, 11, 12, 13, 14, 15, 16 (red). Inputed parameter values: *δ* = 1*/*4 per day; *γ* = 1*/*5.5 per day; *ρ* = 0.5; and infection fatality ratio IFR = 0.9%. Inicial basic reproduction numbers, coefficients of variation and control parameters estimated by Bayesian inference (estimates in Table 1). Fitted curves represent best fitting trajectories and shades are 95% credible intervals generated from 100, 000 posterior samples.

**Fig. 5.**
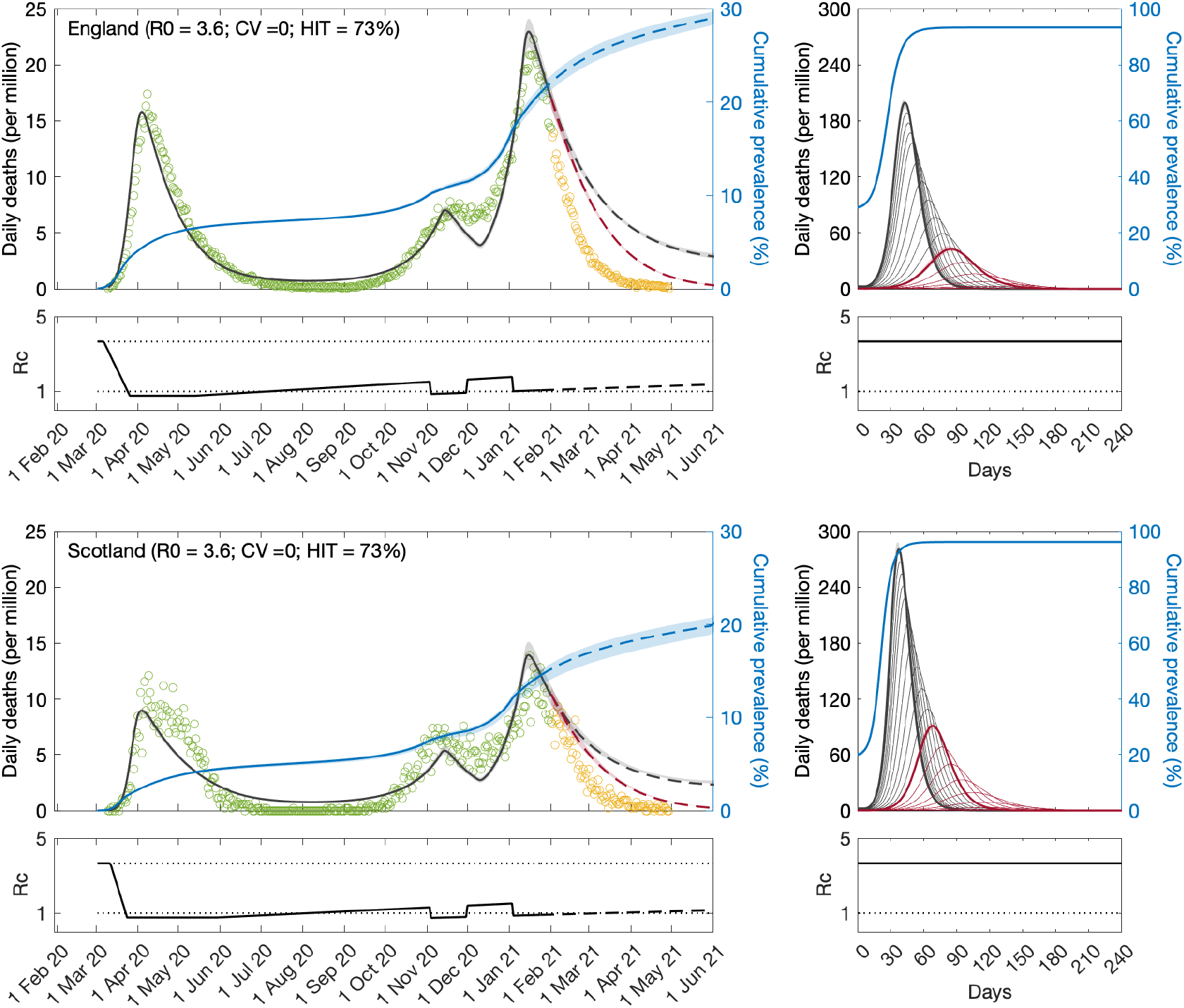
SARS-CoV-2 transmission in England and Scotland under homogeneity. Modelled trajectories of COVID-19 deaths (black and red curves) and cumulative percentage infected (blue). Dots are data for daily reported deaths: fitted (green); posterior to fitted time period (yellow). Basic reproduction numbers under control (ℛ_c_) are displayed on shallow panels underneath the main plots. Left panels represent fitted segments as solid curves and projected scenarios as dashed: without vaccination (black); with a vaccination programme that effectively immunises 8% of the unvaccinated population per month from February onwards (red). Right panels prolong those projections further in time assuming ℛ_c_(*t*) = ℛ_0_ (heavier curves) and explore additional vaccination scenarios (thin curves), from top to bottom (in % of unvaccinated population per month): 0.5, 1, 1.5, 2, 3, 4, 5, 6, 7 (black); 9, 10, 11, 12, 13, 14, 15, 16 (red). Inputed parameter values: *δ* = 1*/*4 per day; *γ* = 1*/*5.5 per day; *ρ* = 0.5; and infection fatality ratio IFR = 0.9%. Inicial basic reproduction numbers and control parameters estimated by Bayesian inference (estimates in Table 1). Fitted curves represent best fitting trajectories and shades are 95% credible intervals generated from 100, 000 posterior samples.

Given the estimated values for ℛ_0_ and *CV* we derive the herd immunity thresholds by natural infection by applying formulas (12) or (19) as appropriate, obtaining ℋ_0_ = 29% in England and ℋ_0_ = 31% in Scotland. If population immunity was to be acquired by random vaccination alone, selection would not play a role and HIT would be given by 1 - 1*/*ℛ_0_, resulting in 71% in England and 73% in Scotland. In reality, both natural infection and vaccination contribute to population immunity, and HIT will be somewhere in between. The homogeneous model suggests ℋ_0_ = 73% in England and ℋ_0_ = 75% in Scotland, in agreement with conventional expectations (Kwok *et al*. 2020).

We then prolong model trajectories (dashed curves) over another 4 months (until 1 June 2021) to compare with data beyond the fitted period (yellow). All models project more deaths than observed, as expected given that the UK initiated a mass vaccination campaign in late 2020 which should start impacting the epidemic by February 2021. To illustrate this we simulate the effective immunisation of 8% of the unvaccinated population per month from February onwards (crude approximation for the UK programme, where 32% were fully vaccinated in the 4-month period February-May) and depict the result by the red dashed curve in the figures. Agreement with data is visually good for the heterogeneous models but insufficient under homogeneity.

The most timely question is perhaps whether achieved population immunity is enough to prevent an exit wave as contact restrictions are lifted. To contribute towards an answers we plot the cumulative number of infections estimated by the model (blue) and find the percentage of the population infected by May 2021 to remain below HIT (more so in Scotland than in England). Hence without vaccination an exit wave might have been expected. To visualise its magnitude we include separate panels on the right where the model is run for 8 months, using as initial conditions the end conditions of the left panels and ℛ_c_(*t*) = ℛ_0_. This is done without vaccination (heavy black) and with vaccination (effective immunisation of 8% per month; heavy red). We explore additional vaccination scenarios (thin curves), from top to bottom (in % of unvaccinated population per month): 0.5, 1, 1.5, 2, 3, 4, 5, 6, 7 (black); 9, 10, 11, 12, 13, 14, 15, 16 (red). We find 1 - 2% effective immunisation by the vaccines per month to be sufficient to prevent the exit wave over the first 8 months, according to the heterogeneous models. This is in stark contrast with the homogeneous scenario, which suggest that only a vaccination programme twice as efficient as the current (e.g., approximately 16% effective immunisation per month) would prevent the exit wave.

In supplementary Information we show that these results are robust to changing IFR (to 0.7% and 1.1%) and including reinfection (with a risk of 0.1 (Hall *et al*. 2021) relative to the average risk of first infection). As expected, assuming a higher IFR results in lower HIT and assuming a lower IFR results in higher HIT. However, when we fit the model with a different IFR, all parameters readjust and the exit wave appears relatively invariant to IFR. The same happens when reinfection is included. The sensitivity analysis is presented for the heterogeneous susceptibility model but replication with heterogeneous connectivity and homogeneity showed similar robustness.

So far we have considered that ℛ_c_(*t*) would always be contained below the original ℛ_0_. Given current concerns that the virus may evolve towards higher transmissibility, such as the emergence of the B.1.1.7 variant (Volz *et al*. 2021; Davies *et al*. 2021; Richard *et al*. 2021), we replicate the above exploration of exit scenarios assuming 50% higher transmissibility (ℛ_c_(*t*) = 1.5 · ℛ_0_) (Fig. 6).

**Fig. 6.**
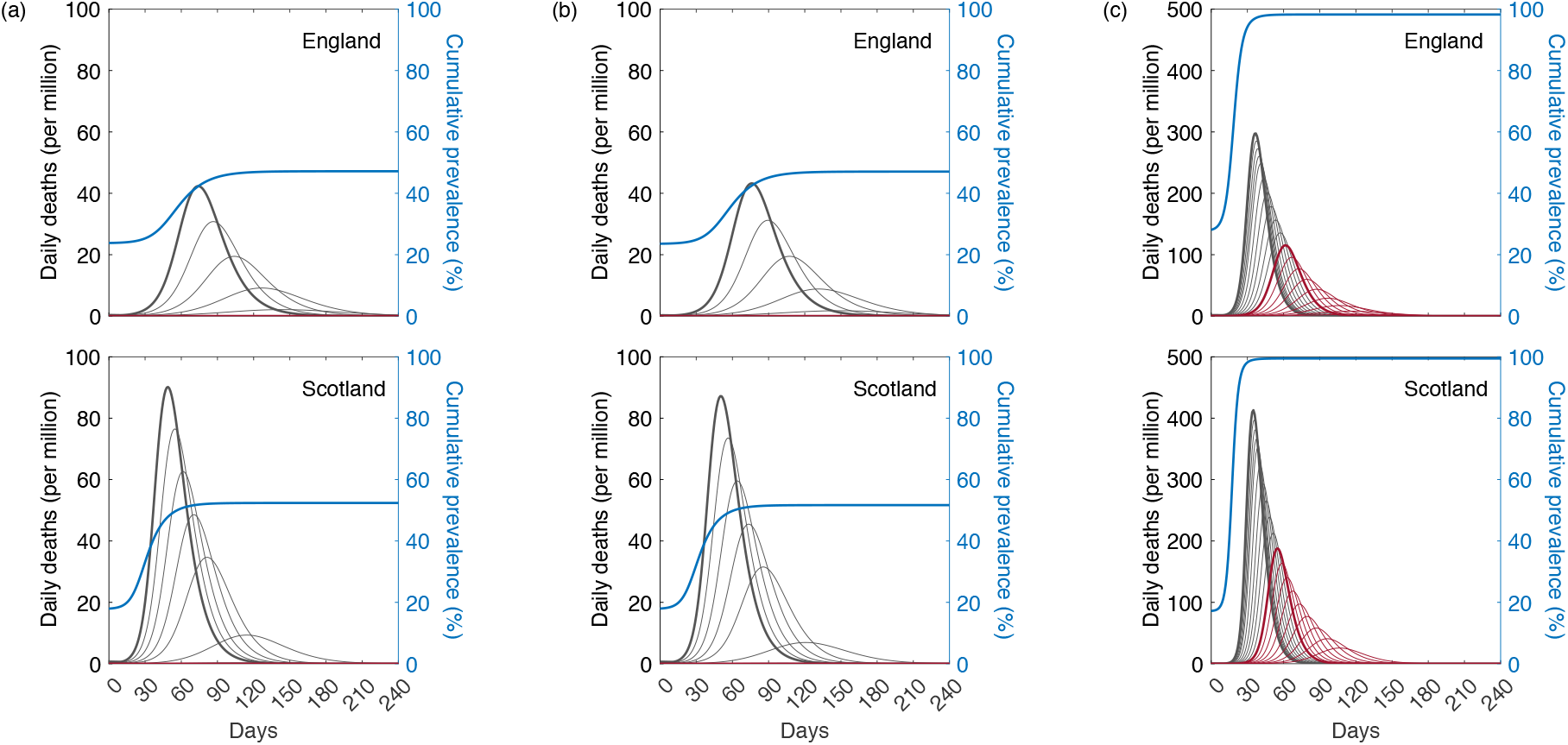
Projected SARS-CoV-2 trajectories with 50% increase in ℛ_0_ in relation to that estimated for the early phase of the pandemic. Assuming ℛ_c_(*t*) = 1.5 ℛ_0_: without vaccination (black heavy curve) and with a vaccination programme that effectively immunises 8% of the unvaccinated population per month from February onwards (red heavy curve). Thin lines explore additional vaccination scenarios, from top to bottom (in % of unvaccinated population per month): 0.5, 1, 1.5, 2, 3, 4, 5, 6, 7 (black); 9, 10, 11, 12, 13, 14, 15, 16 (red). Inputed parameter values: *δ* = 1*/*4 per day; *γ* = 1*/*5.5 per day; *ρ* = 0.5; and infection fatality ratio IFR = 0.9%. (a) Individual variation in susceptibility to infection. (c) Individual variation in exposure to infection. (c) Homogeneous susceptibility and exposure to infection.

### 5.2. Parameter estimation early in the pandemic

In a pandemic it is important to estimate model parameters early when data series are relatively short. To test the suitability of our methods for that task, we apply the fitting procedure to series of COVID-19 deaths in England and Scotland until 1 July 2020, as Europe was just recovering from the first wave (Fig. 7 and Table 2). The results are remarkably similar to those obtained with longer series above (Figs. 3, 4. 5 and Table 1). As before the heterogeneous models are selected as better models than the homogeneous as they have lower AIC. However the difference is not as great as with larger series and in some situations it may not be possible to discriminate.

**Table 2.** Model parameters estimated by Bayesian inference based on daily deaths until 1 July 2020. Model selection based on maximum log-likelihood (LL) and Akaike information criterion (AIC). Best fitting models have lower AIC scores. Infection fatality ratio, IFR = 0.9%. ℋ_0_, calculated from ℛ_0_ and *CV* using formulas (12) or (19), as appropriate.

|  | Heterogeneous susceptibility |  | Heterogeneous connectivity |  | Homogeneous |  |
| --- | --- | --- | --- | --- | --- | --- |
|  | Median | 95% CI | Median | 95% CI | Median | 95% CI |
| *Common parameters* |  |  |  |  |  |  |
| $c_1$ | 0.25 | (0.23, 0.27) | 0.25 | (0.23, 0.27) | 0.23 | (0.22, 0.24) |
| $\eta$ | 15 | (15, 15) | 15 | (15, 15) | 15 | (15, 15) |
| $CV$ | 1.42 | (0.96, 1.81) | 0.99 | (0.43, 1.28) | 0 | — |
| *England* |  |  |  |  |  |  |
| $T_0$ | 0.65 | (0.03, 42.30) | 0.61 | (0.03, 2.77) | 2.27 | (0.23, 4.45) |
| $T_2$ | $1.99 \cdot 10^5$ | $(1.19 \cdot 10^3, 7.00 \cdot 10^7)$ | $7.42 \cdot 10^5$ | $(1.63 \cdot 10^3, 7.95 \cdot 10^7)$ | $9.78 \cdot 10^5$ | $(4.45 \cdot 10^3, 8.08 \cdot 10^7)$ |
| $\mathcal{R}_0$ | 3.43 | (3.31, 3.49) | 3.43 | (3.28, 3.49) | 3.31 | (3.16, 3.48) |
| $\mathcal{H}_0$ | 34% | (24%, 48%) | 34% | (24%, 60%) | 70% | (68%, 71%) |
| *Scotland* |  |  |  |  |  |  |
| $T_0$ | 12.50 | (10.87, 12.98) | 12.48 | (10.78, 12.98) | 12.45 | (10.34, 12.98) |
| $T_2$ | $1.28 \cdot 10^5$ | $(3.55 \cdot 10^2, 6.35 \cdot 10^7)$ | $4.35 \cdot 10^5$ | $(4.09 \cdot 10^2, 6.84 \cdot 10^7)$ | $3.82 \cdot 10^5$ | $(5.83 \cdot 10^2, 7.25 \cdot 10^7)$ |
| $\mathcal{R}_0$ | 3.37 | (3.31, 3.46) | 3.37 | (3.31, 3.46) | 3.37 | (3.30, 3.51) |
| $\mathcal{H}_0$ | 33% | (25%, 47%) | 34% | (24%, 60%) | 70% | (70%, 71%) |
| *Model selection* |  |  |  |  |  |  |
| LL | −900.40 |  | −899.60 |  | −909.89 |  |
| AIC | 1818.80 |  | **1817.20** |  | 1835.80 |  |

**Fig. 7.**
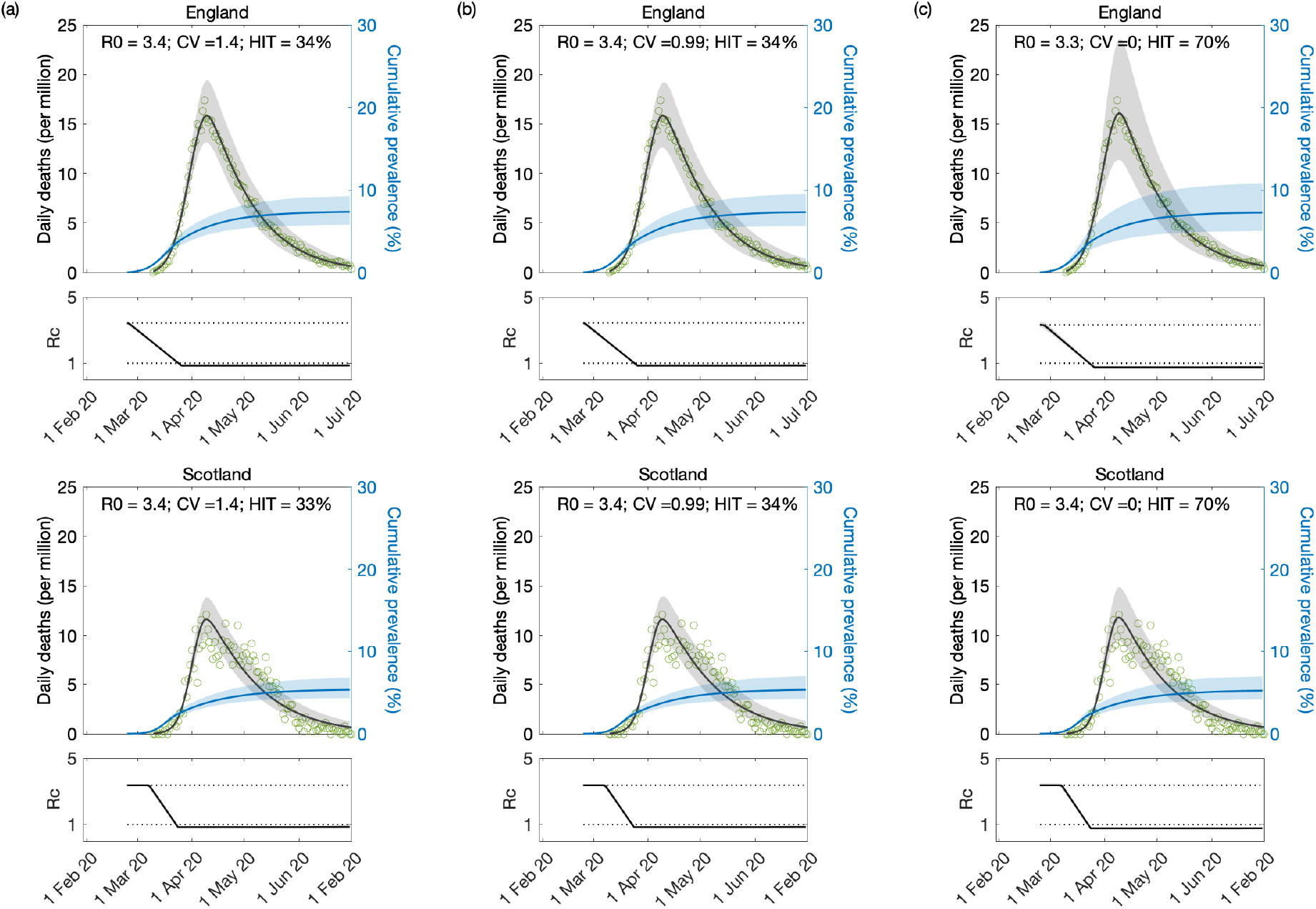
Model-based estimates based on first wave of SARS-CoV-2 pandemic. Modelled trajectories of COVID-19 deaths (black) and cumulative percentage infected (blue). Dots are data for daily reported deaths. Basic reproduction numbers under control (ℛ_c_) are displayed on shallow panels underneath the main plots. Inputed parameter values: *δ* = 1*/*4 per day; *γ* = 1*/*5.5 per day; *ρ* = 0.5; and infection fatality ratio IFR = 0.9%. Inicial basic reproduction numbers, coefficients of variation and control parameters estimated by Bayesian inference (estimates in Table 2). Fitted curves represent best fitting trajectories and shades are 95% credible intervals generated from 100, 000 posterior samples. (a) Individual variation in susceptibility to infection. (c) Individual variation in exposure to infection. (c) Homogeneous susceptibility and exposure to infection.

As an alternative we run the same fits assuming two scenarios for a fixed ramp of contact restriction relaxation out of lockdown (i.e., *T*_2_ fixed). The first is motivated by Table 2, where we find the estimated *T*_2_ to be very large or, equivalently, the ramp to be practically horizontal. Hence we run a scenario where ℛ_c_(*t*) remains strictly horizontal until the end of the fitting period. The support for the heterogeneous models becomes stronger and the estimated parameters (Table 3) remain similar to when *T*_2_ was estimated (Table 2). Second we assume a slope such that if maintained ℛ_c_(*t*) would intersect the original ℛ_0_ in 120 days (i.e., *T*_2_ = 120 days). In this case there is also strong support for the heterogeneous models but the estimated coefficients of variation are larger which is reflected in lower HIT (Table 4).

**Table 3.** Model parameters estimated by Bayesian inference based on daily deaths until 1 July 2020, assuming constant ℛ_c_(*t*) from the first lockdown onwards (i.e. *T*_2_ **→** ∞). Model selection based on maximum log-likelihood (LL) and Akaike information criterion (AIC). Best fitting models have lower AIC scores. Infection fatality ratio, IFR = 0.9%. ℋ_0_, calculated from ℛ_0_ and *CV* using formulas (12) or (19), as appropriate.

|  | Heterogeneous susceptibility |  | Heterogeneous connectivity |  | Homogeneous |  |
| --- | --- | --- | --- | --- | --- | --- |
|  | Median | 95% CI | Median | 95% CI | Median | 95% CI |
| *Common parameters* |  |  |  |  |  |  |
| $c_1$ | 0.25 | (0.23, 0.27) | 0.25 | (0.23, 0.27) | 0.23 | (0.22, 0.24) |
| $\eta$ | 15 | (15, 15) | 15 | (15, 15) | 15 | (15, 15) |
| $CV$ | 1.42 | (0.82, 1.78) | 0.99 | (0.55, 1.30) | 0 | — |
| *England* |  |  |  |  |  |  |
| $T_0$ | 0.58 | (0.03, 2.38) | 0.65 | (0.02, 2.64) | 2.20 | (0.21, 4.38) |
| $\mathcal{R}_0$ | 3.37 | (3.31, 3.46) | 3.43 | (3.28, 3.48) | 3.32 | (3.16, 3.48) |
| $\mathcal{H}_0$ | 34% | (25%, 53%) | 34% | (24%, 54%) | 70% | (68%, 71%) |
| *Scotland* |  |  |  |  |  |  |
| $T_0$ | 12.45 | (10.89, 12.98) | 12.48 | (10.76, 12.98) | 12.43 | (10.31, 12.98) |
| $\mathcal{R}_0$ | 3.37 | (3.31, 3.46) | 3.37 | (3.31, 3.46) | 3.37 | (3.30, 3.51) |
| $\mathcal{H}_0$ | 33% | (25%, 53%) | 34% | (24%, 54%) | 70% | (70%, 71%) |
| *Model selection* |  |  |  |  |  |  |
| LL | −1120.200 |  | −1131.80 |  | −3380.40 |  |
| AIC | **2254.30** |  | 2277.60 |  | 6772.80 |  |

**Table 4.** Model parameters estimated by Bayesian inference based on daily deaths until 1 July 2020, assuming that after the first lockdown ℛ_c_(*t*) begins a gradual return to the baseline ℛ_0_ at a fixed rate (*T*_2_ = 120 days in this case). Model selection based on maximum log-likelihood (LL) and Akaike information criterion (AIC). Best fitting models have lower AIC scores. Infection fatality ratio, IFR = 0.9%. ℋ_0_, calculated from ℛ_0_ and *CV* using formulas (12) or (19), as appropriate.

|  | Heterogeneous susceptibility |  | Heterogeneous connectivity |  | Homogeneous |  |
| --- | --- | --- | --- | --- | --- | --- |
|  | Median | 95% CI | Median | 95% CI | Median | 95% CI |
| *Common parameters* |  |  |  |  |  |  |
| $c_1$ | 0.31 | (0.30, 0.33) | 0.32 | (0.30, 0.35) | 0.24 | (0.23, 0.24) |
| $\eta$ | 15 | (15, 15) | 14 | (13, 15) | 17 | (17, 17) |
| $CV$ | 2.53 | (2.38, 2.66) | 1.86 | (1.70, 2.07) | 0 | — |
| *England* |  |  |  |  |  |  |
| $T_0$ | 0.21 | (0.00, 1.08) | 0.18 | (0.01, 0.97) | 5.48 | (4.02, 6.78) |
| $\mathcal{R}_0$ | 3.44 | (3.38, 3.47) | 3.47 | (3.39, 3.56) | 3.02 | (2.94, 3.11) |
| $\mathcal{H}_0$ | 15% | (14%, 17%) | 15% | (12%, 17%) | 67% | (66%, 68%) |
| *Scotland* |  |  |  |  |  |  |
| $T_0$ | 12.75 | (11.98, 12.99) | 12.79 | (12.13, 12.99) | 12.60 | (11.32, 12.99) |
| $\mathcal{R}_0$ | 3.31 | (3.26, 3.36) | 3.33 | (3.27, 3.41) | 3.23 | (3.19, 3.30) |
| $\mathcal{H}_0$ | 15% | (14%, 17%) | 14% | (12%, 17%) | 69% | (69%, 70%) |
| *Model selection* |  |  |  |  |  |  |
| LL | −951.70 |  | −960.54 |  | −3307.80 |  |
| AIC | **1917.40** |  | 1935.10 |  | 6627.70 |  |

This suggests that in the eventually of future pandemics these models can be used with similar confidence after one or two waves. Although earlier applications might benefit more from knowledge of time-dependent effects of contact restrictions on transmission this information does not appear critical.

### 5.3. Coefficients of variation from contact surveys

Contact patterns provide one of the easiest sources of heterogeneity to study directly. One approach is to use large-scale diary experiments to collect self-reported logs of close or physical contact from study participants. In this section we show data from several of these contact-pattern studies, listed in Table 5. In Fig. 8 we show gamma-fits for the contact distribution of each study on a log scale, along with the CV for each empirical distribution. These empirically measured contact distributions reveal CV between 0.7 and 1.5 (depending on study and setting).

**Table 5.** Contact pattern studies.

|  | Dataset | citation |
| --- | --- | --- |
| 1 | 2008_Mossong_BE | (Mossong *et al.* 2008) |
| 2 | 2008_Mossong_EU | (Mossong *et al.* 2008) |
| 3 | 2008_Mossong_IT | (Mossong *et al.* 2008) |
| 4 | 2008_Mossong_LU | (Mossong *et al.* 2008) |
| 5 | 2008_Mossong_NL | (Mossong *et al.* 2008) |
| 6 | 2008_Mossong_OT | (Mossong *et al.* 2008) |
| 7 | 2008_Mossong_PL | (Mossong *et al.* 2008) |
| 8 | 2008_Mossong_PT | (Mossong *et al.* 2008) |
| 9 | 2008_Mossong_UK | (Mossong *et al.* 2008) |
| 10 | 2009_Hens_BELGIUM | (Hens *et al.* 2009) |
| 11 | 2010_Willem_BELGIUM | (Willem *et al.* 2012) |
| 12 | 2011_Horby_Vietnam | (Horby *et al.* 2011) |
| 13 | 2015_Beraud_France | (Beraud *et al.* 2015) |
| 14 | 2015_Grijalva_Peru | (Grijalva *et al.* 2015) |
| 15 | 2016_Litvinova_Russia | (Litvinova *et al.* 2019) |
| 16 | 2017_Leung_HongKong | (Leung *et al.* 2017) |
| 17 | 2017_Melegaro_Zimbabwe | (Melegaro *et al.* 2017) |
| 18 | 2019_Zhang_China | (Zhang *et al.* 2020) |
| 19 | 2020_Mahikul_Thailand | (Mahikul *et al.* 2020) |
| 20 | 2015_Dodd_ZambiaAndSA | (Dodd *et al.* 2015) |

**Fig. 8.**
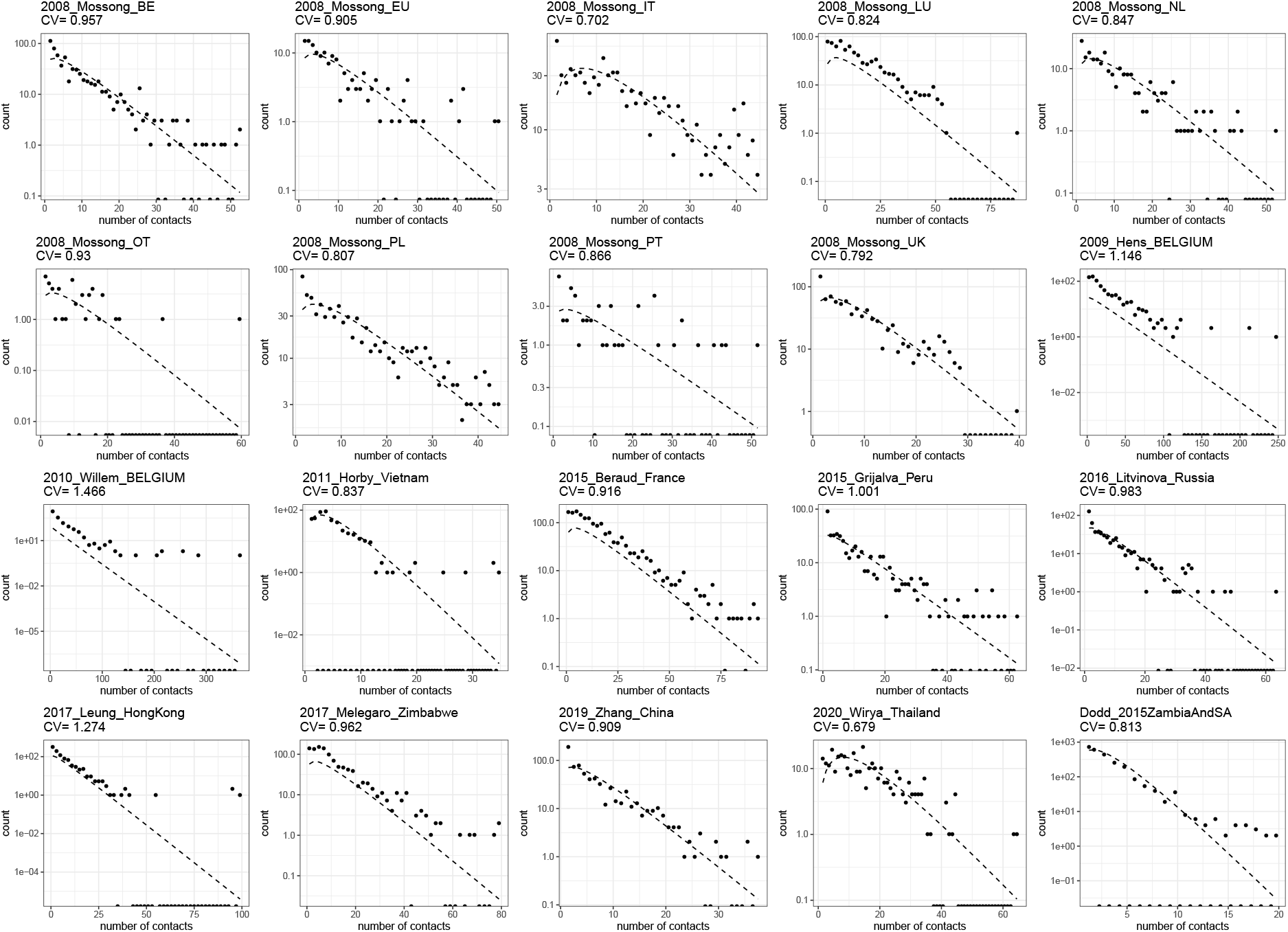
Gamma fits for the included contact surveys.

In addition to the magnitude of individual variation in connectivity, the time scale of that variation is another important determinant of the effect of selection in accelerating the acquisition of population immunity (Tkachenko *et al*. 2021). The referenced studies typically report contact patterns for individuals over a very short (e.g., 1-day) period which is insufficient for assessing persistence of the measured variation. One of the studies (Hens *et al*. 2009) made an extra step and measured contact patterns for each individual on two different days (one weekday and one weekend day). In Fig. 9, we show fits for contact patterns by these two days individually, and for the average. The CV for the contact heterogeneity that persists over the two days is approximately 1.1 (in contrast with the larger 1.4 or 1.6 for each day alone).

**Fig. 9.**
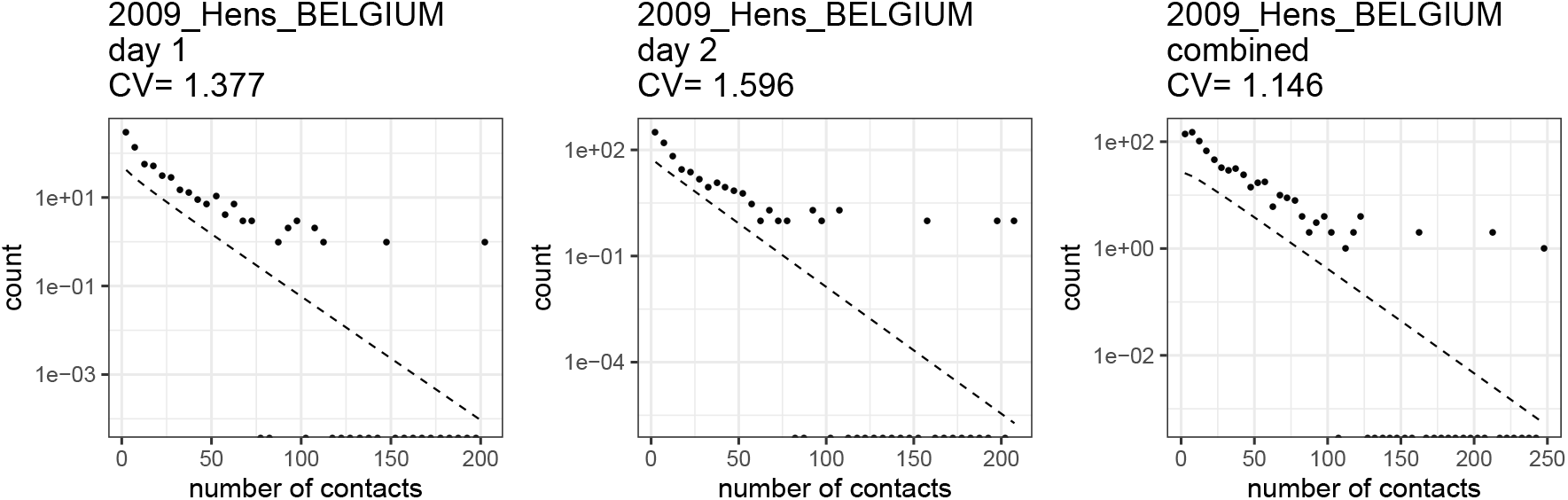
Persistence of contact heterogeneity. When averaged over two separate diary days the contact distribution CV is 17-28% lower.

For a heterogeneous model of an epidemic which unfolds over a timescale like a year, local dynamics are driven by assumptions about heterogeneity in short-term (e.g., week-long) averages in contact patterns, while global dynamics depend on assumptions about persistence of heterogeneity in those short-term averages over the timescale of the simulation. However, these long-term averages are frequently not evaluated directly by contact diary experiments.

One way to estimate persistent contact heterogeneity from below is to bin contact data by age groups. For example, heterogeneity in contact patterns which persists in contact data after binning in 5-year age groups represents population-level heterogeneity in contact patterns that is persistent on multi-year time scales. Of course, this approach only captures heterogeneity mediated by age; i.e., it would only capture the full extent of heterogeneity in contact patterns if there was no within-age-group variation in contact patterns. As such we should expect age-binned contact data to underestimate the level of persistent heterogeneity in contact patterns (Britton *et al*. 2020), perhaps quite substantially. In Fig. 10 we show the effect on CV of binning the data from the studies in Table 5 in 1- and 10-year age groups, respectively. Generally, binning by larger periods tends to reduce heterogeneity substantially.

**Fig. 10.**
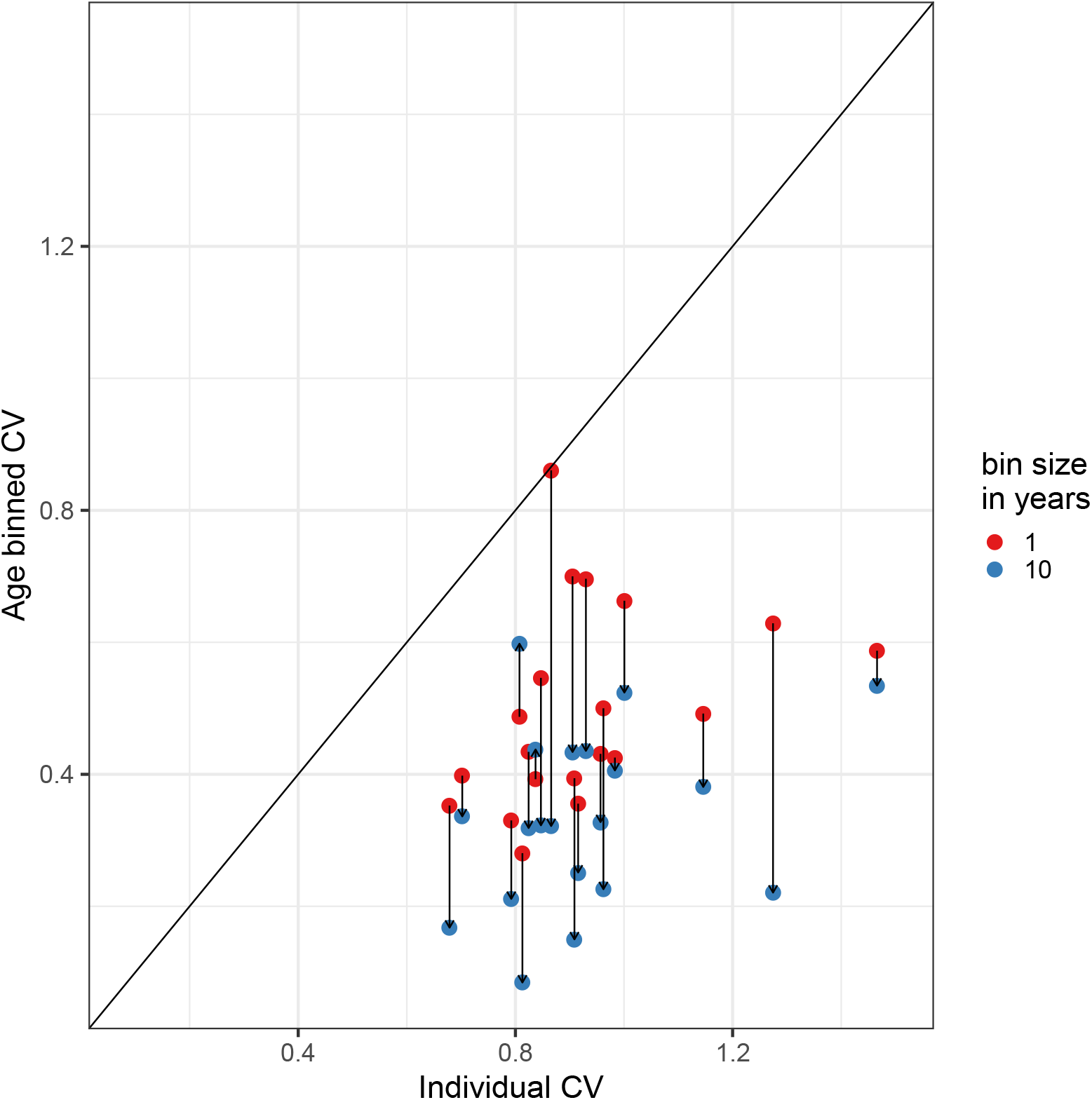
Binning by age.

There have also been detailed studies that specifically measure mobility which suggest higher levels of individual variation in number of contacts (Eubank *et al*. 2004).

An entirely different approach to measure heterogeneity is to trace contacts of infected individuals and count how many secondary infections each has caused. Adam et al (Adam *et al*. 2020) conducted such contact tracing in Hong Kong and estimated a coefficient of variation of 2.5. This is expected to measure more variation as it captures infectiousness as well. The question remains as how persistent this variation is and hence how responsive to selection.

Our model-based inference of persistent heterogeneity based on epidemic trajectories indicates *CV* = 1.1 for England and Scotland (Table 1, heterogeneous connectivity) which appears generally consistent with what is currently available from empirical studies. This level of individual variation lies above that inputed in common epidemic models (whether compartmental or individual-based) that implement contact heterogeneity reduced to 5-year averages and below estimates based on contact tracing which combine the effects of heterogeneity in infectiousness.

## 6. Conclusion

For over a century, mathematical epidemiologists have realised that individual variation in susceptibility and exposure to infection are key determinants of the shape of epidemic curves. Models that underrepresent these forms of variation tend to overpredict epidemic sizes and consequently inflate the effects attributed to control measures. Infectious disease models can include many interacting processes, informed by many data sources, to aid understanding of epidemic dynamics but complexity does not necessarily make them more suitable for predictive purposes. Predictive ability in population dynamics is critically dependent on the complete account of forms of heterogeneity that are under selection. In infectious disease the most impactful selection is that exerted by the force of infection on individual susceptibility and exposure.

With the aim of capturing heterogeneity in full we opted for simple model formalisms with inbuilt distributions of susceptibility or exposure in such a way that we could seek to estimate coefficients of variation by fitting to epidemic curves. We have used the COVID-19 pandemic to test the approach as epidemic trajectories unfold. We estimate final sizes of unmitigated epidemics to be less than half those predicated by studies based on homogeneous models, which is naturally linked to lower herd immunity thresholds. Homogeneous models with realistic vaccination rates also fail to reproduce the low levels of infection and deaths registered over recent months in England and Scotland.

We estimate coefficients of variation consistent with a panoply of empirical studies of contact patterns resulting in herd immunity thresholds by natural infection around 30%, in England and Scotland. Given that vaccination is also contributing to immunisation, herd immunity thresholds are reached with lower infected percentages. Based on forward simulations with a range of vaccination scenarios we conclude HIT is close to being achieved in both nations, with a lower infection burden and greater contribution of vaccination in Scotland. Several countries in Europe and America are in similar situations (Washburne *et al*. 2021) and may be in a comfortable position to redirect vaccines to more susceptible countries.

## Supporting information

Sensitivity analyses

## Data Availability

All data referred to in the manuscript are publicly available.

https://coronavirus.data.gov.uk/

## Acknowledgements

We thank Rodrigo Corder and Antonio Montalbásan for valuable discussions throughout this study.

