## Supplementary material for "Frailty variation models for susceptibility and exposure to SARS-CoV-2": Sensitivity analyses

### Supplementary Information for "Frailty variation models for susceptibility and exposure to SARS-CoV-2": Sensitivity analyses

M. Gabriela M. Gomes

*Department of Mathematics and Statistics, University of Strathclyde, Glasgow, UK, and Centro de Matemática e Aplicações, Faculdade de Ciências e Tecnologia, Universidade Nova de Lisboa, Caparica, Portugal*

Marcelo U. Ferreira

*Institute of Biomedical Sciences, University of São Paulo, São Paulo, Brazil.*

Maria Chikina

*Department of Computational and Systems Biology, University of Pittsburgh, PA, USA.*

Wesley Pegden

*Department of Mathematical Sciences, Carnegie Mellon University, Pittsburgh, PA, USA.*

Ricardo Aguas

*Centre for Tropical Medicine and Global Health, Nuffield Department of Medicine, University of Oxford, Oxford, UK.*

#### Contents:

1. Sensitivity to infection fatality ratio.

2. Sensitivity to reinfection.

#### 1. Sensitivity to infection fatality ratio

##### *1.1. IFR* = $0.7\%$

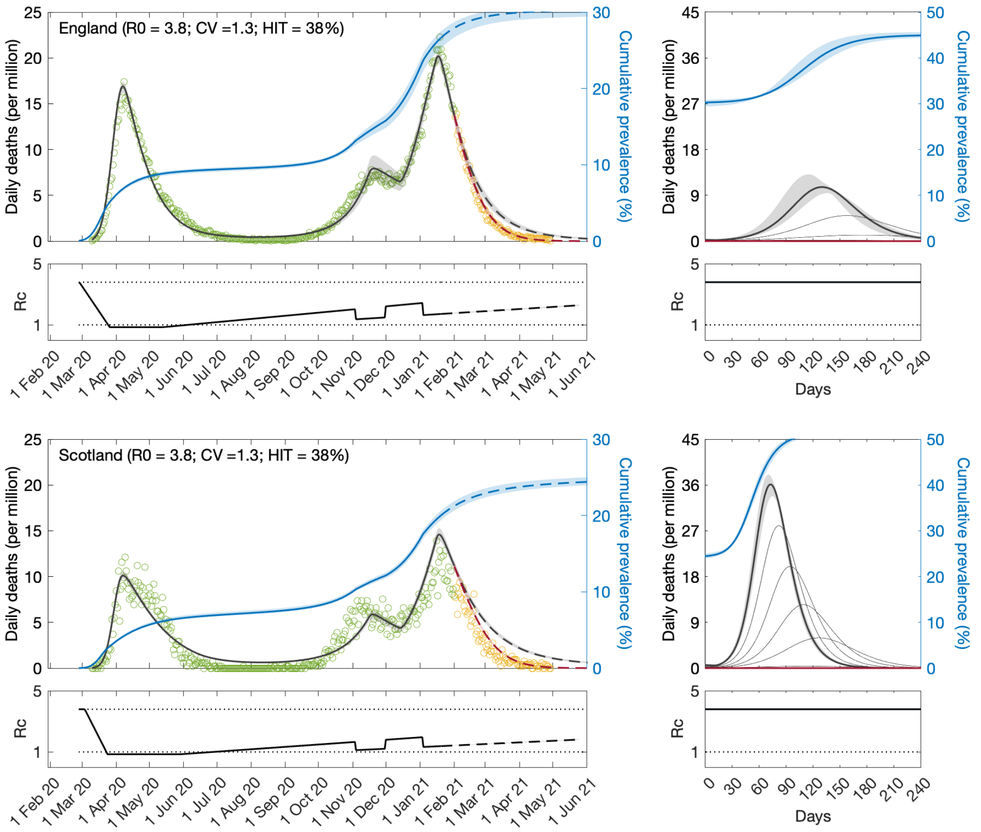


**Fig. S1. SARS-CoV-2 transmission in England and Scotland with individual variation in susceptibility to infection.** Susceptibility factors implemented as gamma distributions. Modelled trajectories of COVID-19 deaths (black and red curves) and cumulative percentage infected (blue). Dots are data for daily reported deaths: fitted (green); posterior to fitted time period (yellow). Basic reproduction numbers under control ($\mathcal{R}_c$) are displayed on shallow panels underneath the main plots. Left panels represent fitted segments as solid curves and projected scenarios as dashed: without vaccination (black); with a vaccination programme that effectively immunises $8\%$ of the unvaccinated population per month from February onwards (red). Right panels prolong those projections further in time assuming $\mathcal{R}_c(t) = \mathcal{R}_0$ (heavier curves) and explore additional vaccination scenarios (thin curves), from top to bottom (in $\%$ of unvaccinated population per month): $0.5, 1, 1.5, 2, 3, 4, 5, 6, 7$ (black); $9, 10, 11, 12, 13, 14, 15, 16$ (red). Inputed parameter values: $\delta = 1/4$ per day; $\gamma = 1/5.5$ per day; $\rho = 0.5$; and infection fatality ratio $\mathrm{IFR} = 0.7\%$. Inicial basic reproduction numbers, coefficients of variation and control parameters estimated by Bayesian inference (estimates in Table S1). Fitted curves represent best fitting trajectories and shades are 95% credible intervals generated from $100,000$ posterior samples.

#### *1.2. IFR* = $1.1\%$

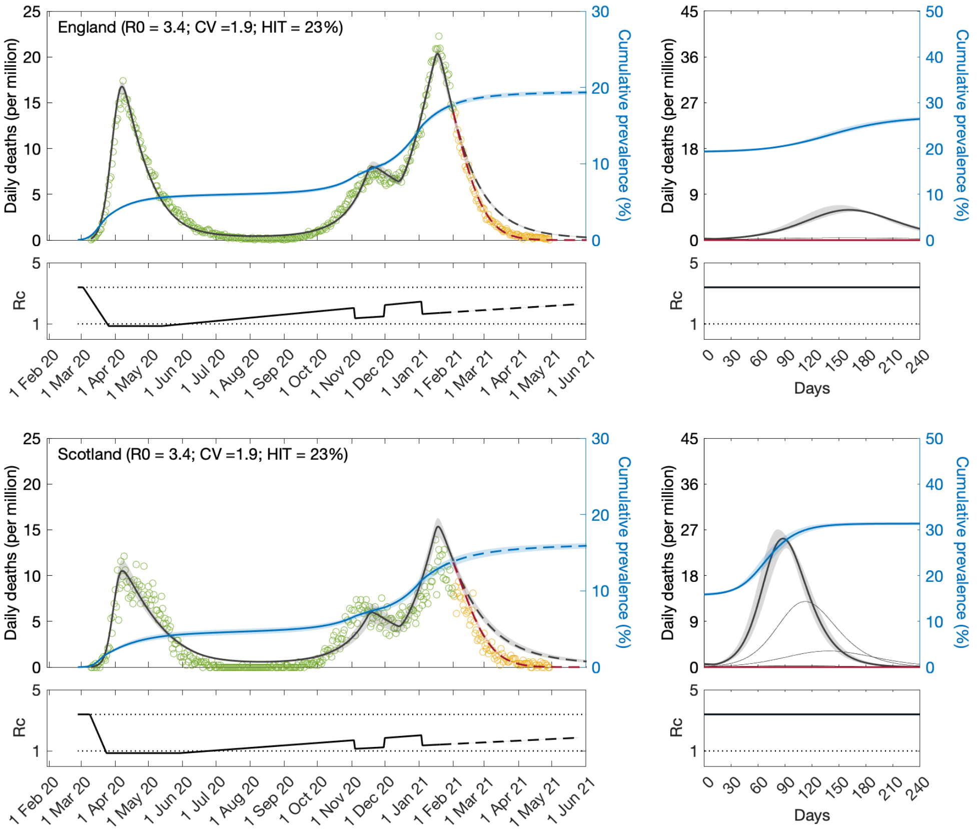


**Fig. S2. SARS-CoV-2 transmission in England and Scotland with individual variation in susceptibility to infection.** Susceptibility factors implemented as gamma distributions. Modelled trajectories of COVID-19 deaths (black and red curves) and cumulative percentage infected (blue). Dots are data for daily reported deaths: fitted (green); posterior to fitted time period (yellow). Basic reproduction numbers under control ($\mathcal{R}_c$) are displayed on shallow panels underneath the main plots. Left panels represent fitted segments as solid curves and projected scenarios as dashed: without vaccination (black); with a vaccination programme that effectively immunises $8\%$ of the unvaccinated population per month from February onwards (red). Right panels prolong those projections further in time assuming $\mathcal{R}_c(t) = \mathcal{R}_0$ (heavier curves) and explore additional vaccination scenarios (thin curves), from top to bottom (in $\%$ of unvaccinated population per month): $0.5, 1, 1.5, 2, 3, 4, 5, 6, 7$ (black); $9, 10, 11, 12, 13, 14, 15, 16$ (red). Inputed parameter values: $\delta = 1/4$ per day; $\gamma = 1/5.5$ per day; $\rho = 0.5$; and infection fatality ratio $\mathrm{IFR} = 1.1\%$. Inicial basic reproduction numbers, coefficients of variation and control parameters estimated by Bayesian inference (estimates in Table S1). Fitted curves represent best fitting trajectories and shades are 95% credible intervals generated from $100,000$ posterior samples.

**Table S1.** Model parameters estimated by Bayesian inference based on daily deaths until 1 February 2021. $\mathcal{H}_0$, calculated from $\mathcal{R}_0$ and $CV$ as appropriate.

| | $IFR = 0.7\%$ | | $IFR = 1.1\%$ | |
|---|---|---|---|---|
| | Median | 95% CI | Median | 95% CI |
| *Common parameters* | | | | |
| $c_1$ | 0.22 | $(0.22, 0.22)$ | 0.25 | $(0.25, 0.26)$ |
| $c_2$ | 0.67 | $(0.67, 0.69)$ | 0.67 | $(0.66, 0.67)$ |
| $\eta$ | 12 | $(12, 12)$ | 12 | $(12, 12)$ |
| $CV$ | 1.33 | $(1.33, 1.33)$ | 1.90 | $(1.88, 1.91)$ |
| *England* | | | | |
| $T_0$ | 0.44 | $(0.08, 0.67)$ | 4.83 | $(4.73, 5.14)$ |
| $T_2$ | 437.74 | $(431.73, 449.72)$ | 372.25 | $(370.24, 374.41)$ |
| $\mathcal{R}_0$ | 3.79 | $(3.77, 3.80)$ | 3.39 | $(3.35, 3.40)$ |
| $\mathcal{H}_0$ | 38% | (38%, 38%) | 23% | (23%, 24%) |
| *Scotland* | | | | |
| $T_0$ | 5.37 | $(5.31, 5.74)$ | 10.92 | $(10.52, 12.01)$ |
| $T_2$ | 578.78 | $(573.28, 587.32)$ | 473.70 | $(466.79, 477.15)$ |
| $\mathcal{R}_0$ | 4.06 | $(4.04, 4.08)$ | 3.60 | $(3.55, 3.64)$ |
| $\mathcal{H}_0$ | 40% | (40%, 40%) | 24% | (24%, 25%) |

#### 2. Sensitivity to reinfection

We formulate reinfection models neglecting individual variation in susceptibility or exposure to the second infection. We set the mean risk of reinfection relative to that of first infection to $\sigma = 0.1$ for illustration purposes and assume that reinfection does not lead to death. In the case of individual variation in susceptibility to the first infection the model is as follows:

$$\dot{S} = -\beta\left[\rho(E+E') + (I+I')\right]\left(\frac{S}{N}\right)^{1+CV^2}, \tag{1}$$

$$\dot{E} = \beta\left[\rho(E+E') + (I+I')\right]\left(\frac{S}{N}\right)^{1+CV^2} - \delta E, \tag{2}$$

$$\dot{I} = \delta E - \gamma I, \tag{3}$$

$$\dot{S}' = (1-\phi)\gamma I - \sigma\beta\left[\rho(E+E') + (I+I')\right]\frac{S'}{N}, \tag{4}$$

$$\dot{E}' = \sigma\beta\left[\rho(E+E') + (I+I')\right]\frac{S'}{N} - \delta E', \tag{5}$$

$$\dot{I}' = \delta E' - \gamma I', \tag{6}$$

with basic reproduction number:

$$\mathcal{R}_0 = \beta\left(\frac{\rho}{\delta} + \frac{1}{\gamma}\right). \tag{7}$$

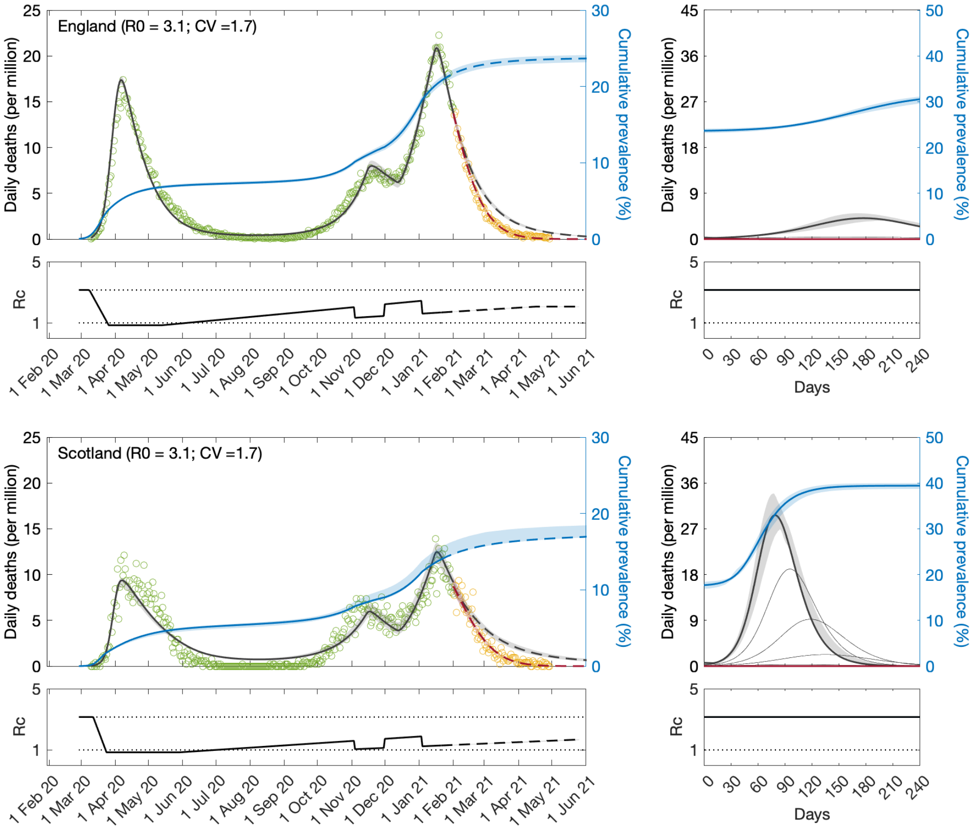


**Fig. S3.** **SARS-CoV-2 transmission in England and Scotland with reinfection and individual variation in susceptibility to the first infection.** Susceptibility factors implemented as gamma distributions. Modelled trajectories of COVID-19 deaths (black and red curves) and cumulative percentage infected (blue). Dots are data for daily reported deaths: fitted (green); posterior to fitted time period (yellow). Basic reproduction numbers under control ($\mathcal{R}_c$) are displayed on shallow panels underneath the main plots. Left panels represent fitted segments as solid curves and projected scenarios as dashed: without vaccination (black); with a vaccination programme that effectively immunises $8\%$ of the unvaccinated population per month from February onwards (red). Right panels prolong those projections further in time assuming $\mathcal{R}_c(t) = \mathcal{R}_0$ (heavier curves) and explore additional vaccination scenarios (thin curves), from top to bottom (in $\%$ of unvaccinated population per month): $0.5$, $1$, $1.5$, $2$, $3$, $4$, $5$, $6$, $7$ (black); $9$, $10$, $11$, $12$, $13$, $14$, $15$, $16$ (red). Inputed parameter values: $\delta = 1/4$ per day; $\gamma = 1/5.5$ per day; $\rho = 0.5$; infection fatality ratio $\mathrm{IFR} = 0.9\%$; and reinfection factor $\sigma = 0.1$. Inicial basic reproduction numbers, coefficients of variation and control parameters estimated by Bayesian inference (estimates in Table S2). Fitted curves represent best fitting trajectories and shades are 95% credible intervals generated from $100,000$ posterior samples.

**Table S2.** Model parameters estimated by Bayesian inference based on daily deaths until 1 February 2021. Individual variation in susceptibility to the first infection. Reinfection factor: $\sigma = 0.1$.

| | Median | 95% CI |
|---|---|---|
| *Common parameters* | | |
| $c_1$ | 0.27 | $(0.27, 0.27)$ |
| $c_2$ | 0.66 | $(0.65, 0.69)$ |
| $\eta$ | 11 | $(11, 11)$ |
| $CV$ | 1.67 | $(1.66, 1.70)$ |
| *England* | | |
| $T_0$ | 9.46 | $(9.38, 9.80)$ |
| $T_2$ | 339.12 | $(335.96, 341.52)$ |
| $\mathcal{R}_0$ | 3.15 | $(3.12, 3.16)$ |
| *Scotland* | | |
| $T_0$ | 2.94 | $(1.45, 3.00)$ |
| $T_2$ | 485.24 | $(480.66, 528.49)$ |
| $\mathcal{R}_0$ | 3.45 | $(3.42, 3.47)$ |
